# The Detection of COVID-19 in Chest X-Rays Using Ensemble CNN Techniques

**DOI:** 10.1101/2022.11.29.22282856

**Authors:** Domantas Kuzinkovas, Sandhya Clement

## Abstract

Advances in the field of image classification using convolutional neural networks (CNNs) have greatly improved the accuracy of medical image diagnosis by radiologists. Numerous research groups have applied CNN methods to diagnose respiratory illnesses from chest x-rays, and have extended this work to prove the feasibility of rapidly diagnosing COVID-19 to high degrees of accuracy. One issue in previous research has been the use of datasets containing only a few hundred images of chest x-rays containing COVID-19, causing CNNs to overfit the image data. This leads to a lower accuracy when the model attempts to classify new images, as would be clinically expected of it. In this work, we present a model trained on the COVID-QU-Ex dataset, overall containing 33,920 chest x-ray images, with an equal share of COVID-19, Non-COVID pneumonia, and Normal images. The model itself is an ensemble of pre-trained CNNs (ResNet50, VGG19, VGG16) and GLCM textural features. It achieved a 98.34% binary classification accuracy (COVID-19/no COVID-19) on a balanced test dataset of 6581 chest x-rays, and 94.68% for distinguishing between COVID-19, Non-COVID pneumonia and normal chest x-rays. Also, we herein discuss the effects of dataset size, demonstrating that a 98.82% 3-class accuracy can be achieved using the model if the training dataset only contains a few thousand images, but that generalisability of the model suffers with such small datasets.

## I. Introduction

Rapid diagnosis of COVID-19 in hospitals is vital for ensuring that patients with respiratory symptoms are triaged swiftly and receive the correct treatment. The state of the art for confirming a suspected COVID-19 case is the use of Reverse Transcriptase Polymerase Chain Reaction (RT-PCR). However, the process of obtaining PCR results is slow, and some studies have found it to only be sensitive to about 90.7% [1]. One alternative is to perform a chest x-ray, which takes 10 minutes or less, and then use a deep learning model to diagnose the patient, taking milliseconds. Deep learning models are also typically more sensitive in detecting diseases from medical images than radiologists [2], and as this work will show, can be more sensitive to detecting COVID-19 than a PCR.

Ever since the beginning of the COVID-19 pandemic, deep learning approaches for the detection of coronavirus pneumonia in chest x-rays, and its distinction from an alternative pneumonia, became of great interest to the research community. Several groups have presented promising results using variations of convolutional neural network (CNN) based image recognition models [3]-[10].

Some models utilise only a single CNN for classification such as in Wang et al. (2020), where a custom 89-layer CNN named COVID-Net was developed [3]. The group obtained a 93.3% accuracy for distinguishing between chest X-rays containing COVID-19 pneumonia, other pneumonia or no condition. However, due to the novelty of the pandemic at the time, only 358 of the total 13,975 X-rays the group obtained were examples of a COVID-19 infection, making effective training of the model difficult due to the image class bias. It is not exclusively required to develop a custom CNN to perform medical image diagnoses, however. Instead, a process known as transfer learning can be used, in which the feature extraction ability learned by a CNN trained on a different dataset can be transferred to assist in classifying images from a different application [4]. Zouch et al. (2022) employed a CNN transfer learning approach, comparing the performance of the ResNet50 and VGG19 CNNs pretrained on the ImageNet dataset [5]. Between the two models, VGG19 had superior performance with a 99.35% binary classification accuracy (COVID-19/No COVID-19) compared to the 96.77% accuracy for ResNet50. However, due to the small and unbalanced dataset size of 112 COVID-19 and 747 Non-COVID-19 chest X-rays, overfitting of the dataset may have feasibly occurred. Despite this, the study conveys the effectiveness of transfer learning in a medical image classification application, proving that CNNs do not have to be built from scratch to obtain high classification accuracies.

CNNs do not have to be used for the classification step, but can alternatively be utilised as feature extraction tools before passing these features to other types of classifiers [6]-[10]. Sethy et al. (2020) achieved a 4-class accuracy of 95.33% by combining ResNet50 with a Support Vector Machine (SVM) classifier [7]. Karim et al. (2022) combined the features extracted using AlexNet with several types of machine learning classifiers and obtained a maximum 3-class accuracy of 98.01% by transferring these features to a Naïve-Bayes classifier [9].

Models with more complex construction have been shown to achieve very high accuracies on medium-sized datasets. Notably, the methods of Mostafiz et al. (2022) included watershed segmentation, GLCM/Wavelet feature extraction, ResNet50 for feature extraction, feature selection using Maximum Relevance Minimum Redundancy (mRMR) and Recursive Feature Elimination (RFE), and a final Random Forest Classifier for classification [6]. Due to the high number of optimisation steps, the accuracy obtained was 98.48% for a 4-class classification (COVID-19, Bacterial Pneumonia, Viral Pneumonia and Normal). Their dataset was a combination of previous existing datasets, with 4809 chest X-rays, 790 of which contained COVID-19 infections.

Another example of this is in Toğaçar et al. (2020), where three CNNs were used to extract features from 5849 chest X-rays of normal and non-COVID pneumonia cases [10]. An mRMR feature selection algorithm was used to determine the most important features. The group concluded that the best configuration involved selecting 100 features from each CNN before passing them to a Linear Discriminant Analysis (LDA) classifier. This configuration obtained a 99.41% binary classification accuracy. The benefit of such a system is that the classification outcome is a collaborative effort of several CNNs of different architectures, meaning that features missed by one CNN may be accounted for by another CNN. This makes it more difficult for classification errors to occur [10].

The discussed literature provides great insight into the variety of viable models for classification of chest x-ray images. However, due to the novelty of COVID-19 at the time, the number of COVID-19 chest x-rays utilised by the studies did not exceed 3616 [9], and most only had below 1000. Furthermore, many of these studies do not explore the generalisability of their models on datasets external to their training datasets, preventing knowledge of their clinical effectiveness. In the present study, we train our model on a dataset of almost 33,000 total images, a third being of COVID-19 infections, and we show that this has a marked improvement in generalisability performance compared to training on a smaller dataset. We also expand on previous works by combining features from multiple CNNs with GLCM features, and by exploring the relative benefit of RF, LDA, LR and ANN classifiers to classify the combined features.

## II. Methods

### A. Datasets

The dataset used for training and evaluation of the model was the COVID-QU-Ex dataset developed by researchers at Qatar University and University of Dhaka [11]-[13]. This is a dataset of 33,920 chest x-rays, of which 11,956 contain a COVID-19 infection, 11,263 contain bacterial or viral infections and 10,701 are normal. This dataset was chosen for its large size, and its balanced nature, which help to tackle overfitting and biased learning respectively. Prior to using the dataset, it was cleaned by removing x-ray images with excessively high or low contrast. This lowered the numbers of images in the training set from 21715 to 21102, the validation set from 5417 to 5274, and the test set from 6788 to 6581. Examples of x-rays from each class can be found in Fig. 1.

**Fig. 1.**
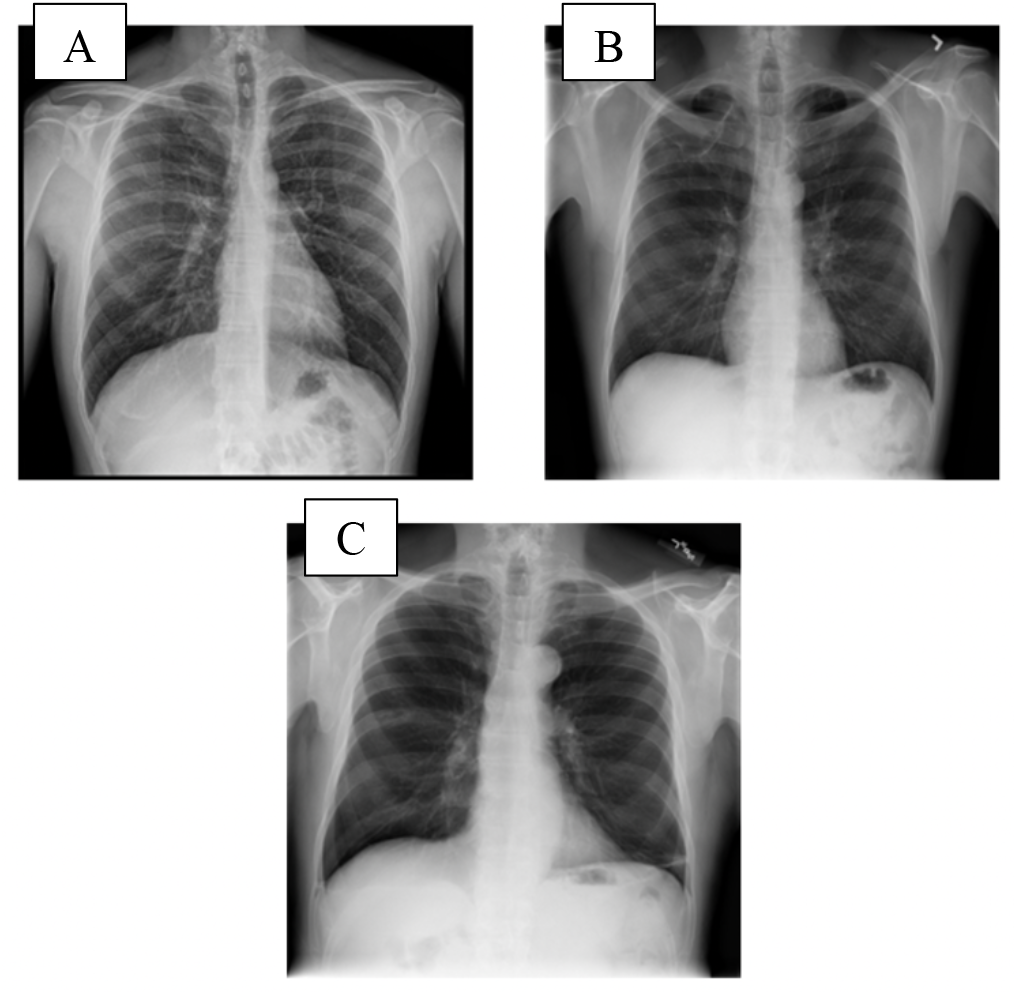
Examples of COVID-19 (A), Non-COVID Pneumonia (B) and Normal (C) chest x-ray images from the COVID-QU-Ex dataset [11].

Many of the other research works discussed in the previous section used far smaller datasets, some with only a few hundred x-ray images of each of COVID-19 infections, other pneumonia infections, and no infections. Many of these papers also claim high (>98%) accuracies in the classification of the chest x-rays. Great care must be taken when using smaller datasets to avoid the issue of overfitting. Dataset overfitting is the phenomenon whereby a classifier performs poorly on datasets which were not used to train the classifier. It is often the result of not having enough training images to teach the classifier to extract generalised features from images of each class. Instead, it learns to extract features that are specific to the dataset it was given, and therefore performs worse when it cannot find these features in other datasets. This issue is far from trivial, since clinical use of such a chest x-ray classification system requires that it is robust and accurate, no matter the x-ray image’s source or properties.

To explore the issue of overfitting in medical image literature, a smaller dataset of 4809 chest x-ray images was also obtained from Mostafiz et al. (2022) [6][14]. It contains 790 cases of COVID-19, 2519 cases of bacterial or viral pneumonia, and 1500 normal cases. It is actually composed of 3 datasets: COVID-19 images from Cohen et al. (2020) [15] and Dadario (2020) [16], and normal and pneumonia images from Kermany et al. (2018) [17]. A summary of the dataset statistics can be found in Table 1.

**TABLE 1:** Dataset sample numbers for COVID-QU-Ex and Mostafiz et al. (2022) DATASETS.

| Dataset and Classes | Train | Val | Test | Total |
| --- | --- | --- | --- | --- |
| **COVID-QU-Ex [12]** |  |  |  |  |
| COVID-19 | 7290 | 1826 | 2264 | **11380** |
| Non-COVID Pneumonia | 7082 | 1762 | 2204 | **11048** |
| Normal | 6730 | 1686 | 2113 | **10529** |
| **Total** | **21102** | **5274** | **6581** | **32957** |
| **Mostafiz et al. (2022) [14]** |  |  |  |  |
| COVID-19 | 442 | 111 | 237 | **790** |
| Non-COVID Pneumonia | 1410 | 353 | 756 | **2519** |
| Normal | 840 | 210 | 450 | **1500** |
| **Total** | **2692** | **674** | **1443** | **4809** |

### B. Model Overview

The model is an adaptation of that used in Toğaçar et al. (2020) [10] to the application of COVID-19 detection, and is illustrated in Fig. 3. In general, it involves the combination of features extracted using several CNNs, in this case ResNet50, VGG19 and VGG16, and extracted GLCM features. The CNN features for one image consist of a vector of 1024 output values from the final Dense layer of each CNN, the layer immediately before the classification into the 3 image classes.

The 80 GLCM features extracted from the Grey-Level Co-occurrence Matrix give details about textural features in the image, such as pixel contrast, energy, homogeneity and correlation.

The 1024-value feature vectors from the CNNs are then shortened to vectors of only the 160 most important features for correctly classifying the chest x-ray. For GLCM features, 80 of 144 were selected. The selection criteria were determined by an mRMR (Minimum Redundancy Maximum Relevance) algorithm, available as a library in Python [18]. The purpose of performing this feature selection is to minimise computation time and to prevent less irrelevant features from causing incorrect classifications.

Once the feature selection is performed, the 560 total features were horizontally concatenated and this vector was passed to one of multiple traditional classifiers such as a Random Forest Classifier, to fit the classification model. A separate test dataset was then used to evaluate the performance various model combinations and configurations.

### C. Convolutional Neural Networks (CNNs)

CNNs by themselves are able to relatively accurately classify medical images. The combined efforts of multiple CNNs, however, can allow for superior results to any of the individual CNNs. The current model utilised 3 CNNs loaded with weights that were pretrained on the ImageNet dataset [20]. During this pretraining, the CNNs learned how to extract various features such as edges, patterns and textures from images of objects, including animals, vehicles and food items.

The ability to extract such features from non-medical images can be “transferred” to a medical image classification application, known as transfer learning [4]. The 3 pretrained CNNs used were ResNet50, VGG19 and VGG16, each of which were prepared in Python using Tensorflow and Keras [19]. The preparation involved removing their classification layers, and adding a Dense-1024 layer, followed by a dropout layer, another Dense-1024 layer, and a Dense-3 layer as the final classification layer, as shown in Fig. 2. The dropout layer was added as an additional way to combat overfitting during the training of the CNNs. A value of 30% of randomly dropped input neurons was used. The Dense-3 layer was added to the CNN just for training of the layer weights, with each node representing one of either COVID-19, Non-COVID, or Normal classes. The layer was removed when the models were later used for feature extraction (where the final layer was Dense-1024), and the dropout was inactive during this extraction stage.

**Fig. 2.**
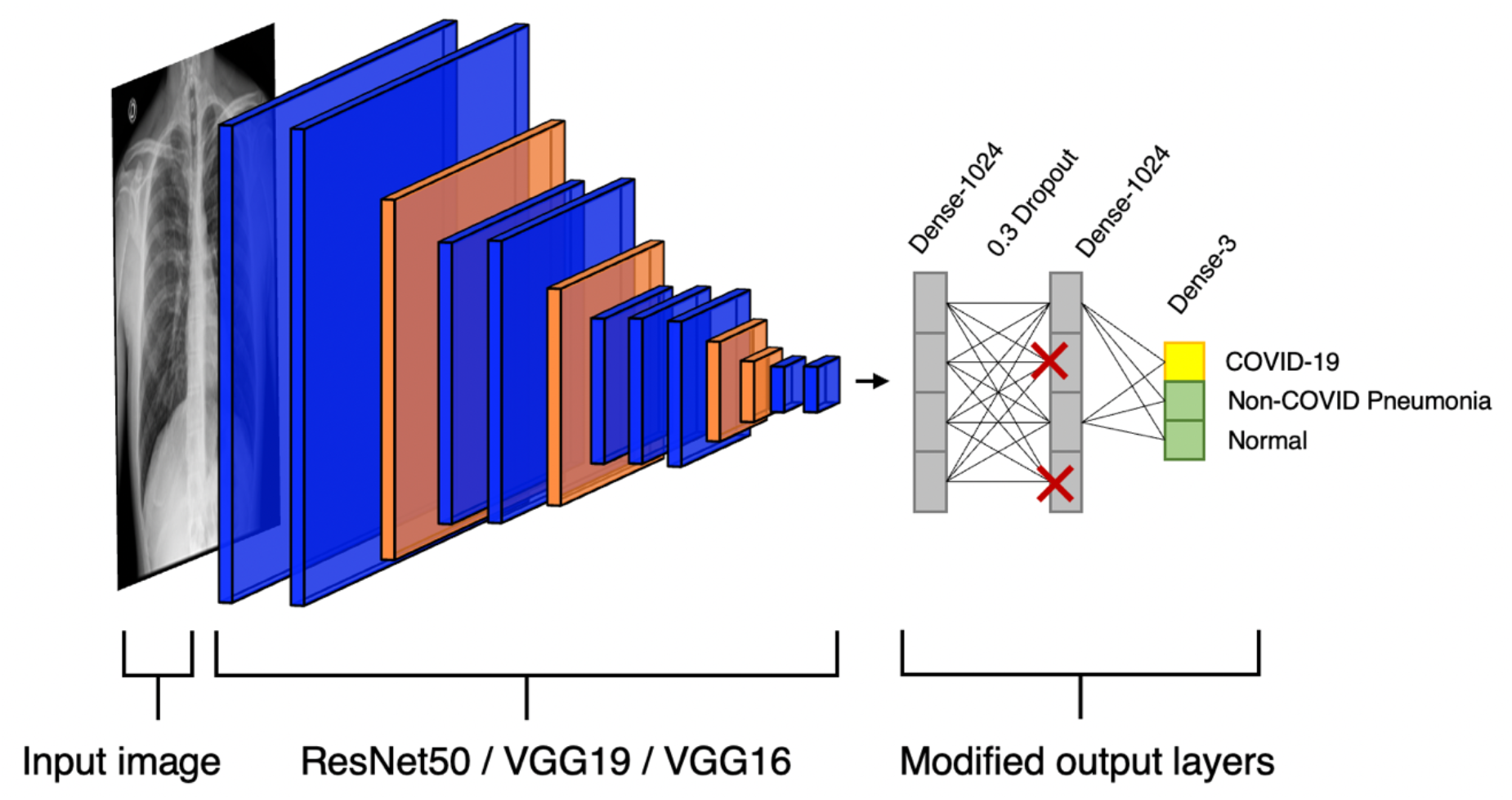
Schematic of how pretrained CNNs were modified to classify chest x-rays. Later, the CNNs were used for feature extraction purposes, whereby the Dense-3 layer was removed and the final Dense-1024 layer (without dropout) was used to provide 1024 features for classification.

**Fig. 3.**
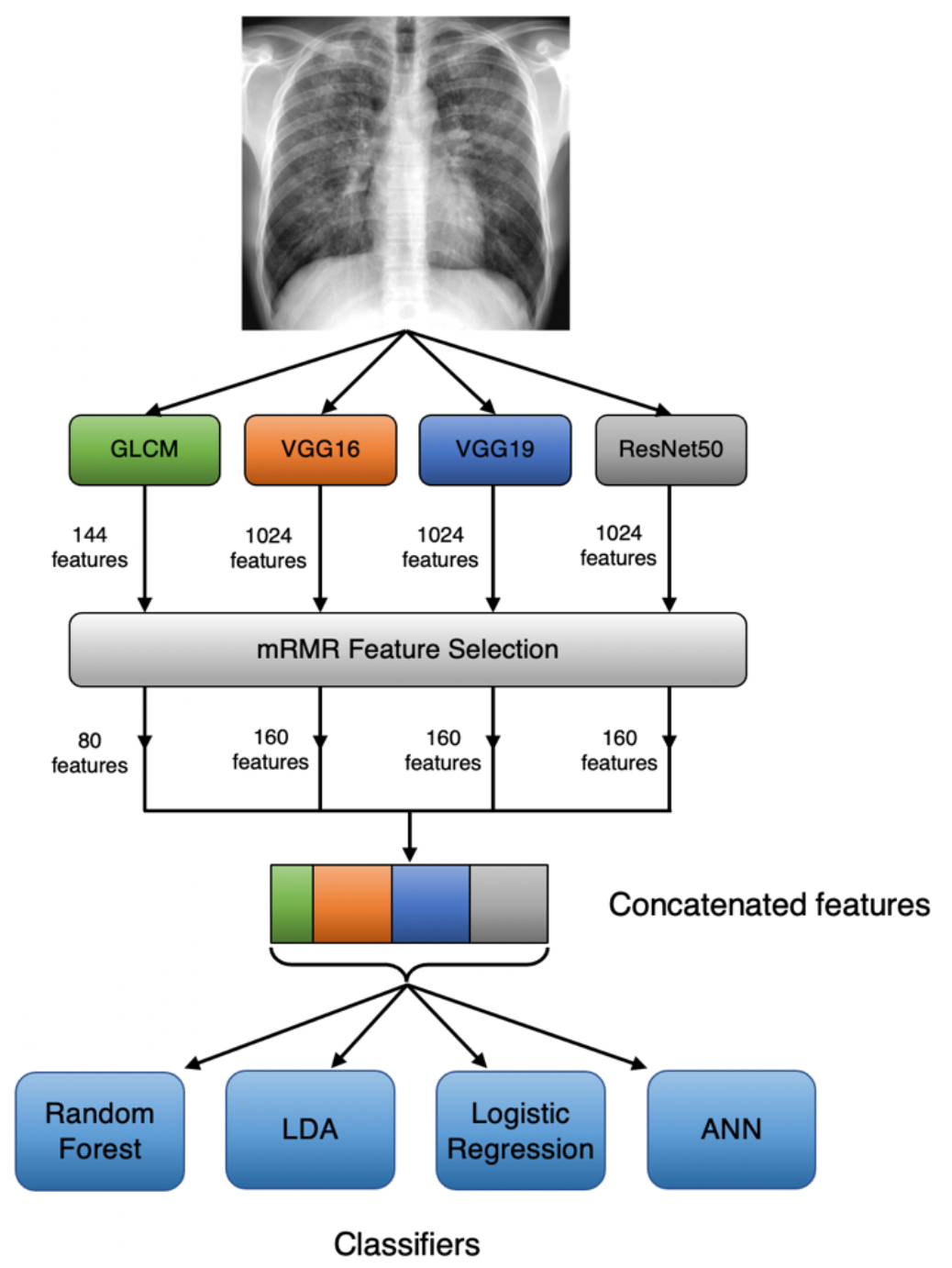
Proposed ensemble CNN and GLCM chest x-ray classification model.

Before training, the base layers of the model (with weights trained on ImageNet) were frozen such that only the last 3 layers were trainable. This is common practice in deep learning, aiming to reduce computation time. Rather than training each CNN for a fixed number of epochs, a learning rate reduction and early stopping procedure was used, to strategically shift the weights towards convergence. The learning rate was first set to 0.001 and the validation loss was monitored. If the validation loss did not improve (decrease) for 3 epochs, known as the “patience” in Keras, the learning rate was set to 0.1 times the previous value. The minimum learning rate was set to 1e-6. At any stage, if there was no improvement in the validation loss for 6 epochs, the training process was exited. This training procedure was performed with the Adam optimiser, and a batch size of 32. Training was performed in a Jupyter notebook on an Apple MacBook Pro with M1 Max using its GPU.

### D. Grey-Level Co-occurrence Matrix Features

A Grey-Level Co-occurrence Matrix (GLCM), first proposed by Haralick et al. (1973), is a compact method of expressing the number of times a certain pair of pixels appears along a particular direction in an image, and at a particular distance away from each other [21]. Its purpose is to allow for the computation of textural features within the image, such as its contrast and homogeneity. Given a greyscale image with 256 distinct levels of grey, its co-occurrence matrix *P*_*i,j*_ will be of size 256 × 256. Assuming computation in the horizontal 0° direction and distance = 1, each value (*i, j*) in the matrix equals the number of times the pixel value pair *i, j* appeared in the original image horizontally and with *j* directly adjacent to *i*, as illustrated in Fig. 4.

**Fig. 4.**
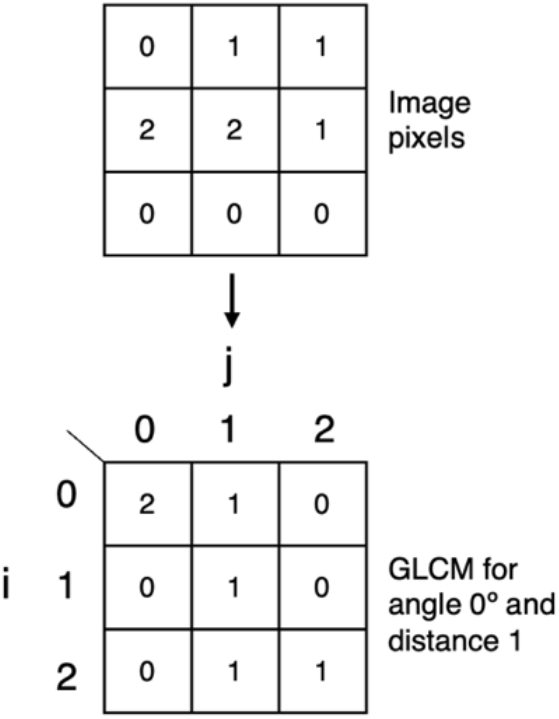
GLCM transformation on 3×3 arrangement of pixels with values 0-2. Performed on 0º angle and pixel distance 1.

Using a GLCM, several textural image properties can be computed. A summary of the equations for calculating various image properties have been outlined in Table 1. The feature extraction from the GLCM was performed using the Sci-Kit Image Python library [22]. Each of the six GLCM image properties in Table 2 were computed. These were computed in 8 directions and 3 distances in order to obtain as much information from each chest x-ray image as possible. This amounts to a total of 144 features (6 categories x 8 directions x 3 distances), where each “feature” is defined as a numerical value of one of the GLCM properties for a particular direction and distance.

**TABLE 2:**
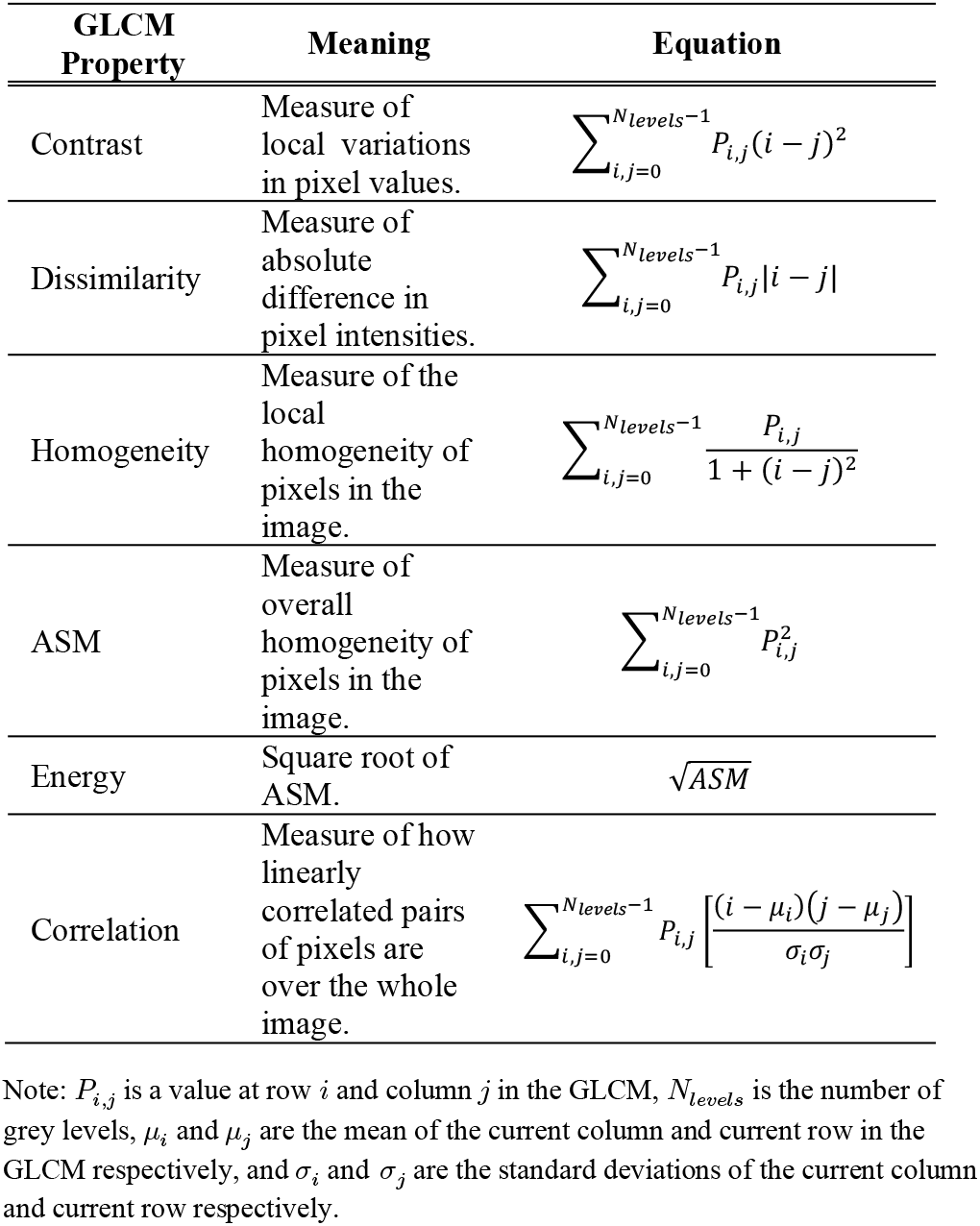
GLCM properties and their definitions.

### E. mRMR Feature Selection

The Minimum Redundancy Maximum Relevance algorithm proposed by Ding and Peng (2005) aims to select features with the most importance to classification when using traditional (non-CNN) classifiers [23][24]. Removing irrelevant features allows for quicker computation time when fitting the model to a traditional classifier, as well as more accurate results as there are less features to consider and therefore fewer chances of model confusions [25]. Their algorithm iteratively cycles through the features and extracts the most relevant and least redundant feature at each iteration. On the first iteration, the feature with the highest F-test statistic is selected. On subsequent iterations, the criterion for selection is a feature’s F-statistic divided by its average Pearson correlation to all features selected on previous iterations as in Equation 1 below. This is known as the F-test Correlation Quotient (FCQ).

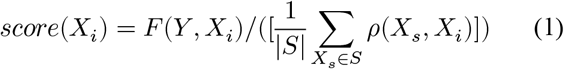

*X*_*i*_ is the feature to be selected at iteration *i, F* (*Y, X*_*i*_) is the F-test statistic of the feature with respect to its corresponding class label *Y, S* is the set of previously selected features, and *ρ*(*X*_*s*_, *X*_*i*_) is the Pearson correlation coefficient of the feature to each of the previously selected features, *X*_*s*_.

There are many variations of this criterion depending on the specific use case. For example, if the classifier to be used on the feature set is a Random Forest Classifier, the F-test relevance criterion can be replaced with one derived from the decision tree algorithm of the classifier, known as the Gini feature importance, in order to further improve the relevance of the selected features [26]. The resulting mRMR feature selection algorithm is then known as RFCQ (Random Forest Correlation Quotient).

For the present study, RFCQ was used. The number of iterations, and hence the number of features selected, was set to 160 for each of the feature sets generated by the CNN models, and to 80 for the GLCM feature set. These were the values that gave the best performance when considering computation time.

### F. Classification Process

In a secondary “training” process, a Python program was created to extract feature sets from each of the chest x-rays images in each of the COVID-QU-Ex and Mostafiz et al. training datasets. Each of the 3 trained CNNs provides 1024 features directly from their Dense-1024 layer for each image. The 144 GLCM features are also extracted in this process. Once this is complete, the features from each source are processed by the mRMR algorithm, taking only the 160 most important features from each CNN feature set, and 80 from the GLCM feature set. The resulting features are then concatenated to form a final feature set of 560 features for each of the chest x-ray images in the training dataset.

The resulting matrix of size *n*_*images*_ × 560 was then passed to one of 4 classifiers, each of which is outlined below:

#### 1) Random Forest Classifier (RF)

A Random Forest Classifier involves a series of individual decision tree classifiers (estimators), that individually attempt to classify randomly selected feature samples [27]. While these estimators may make errors, the majority vote of each of the many estimators gives a much more accurate prediction, leading to the success of RFs. In the current study, an RF implemented in the Sci-kit Learn Python library, with 200 estimators.

#### 2) Linear Discriminant Analysis (LDA)

LDA works by grouping features such that variance between classes is maximised and variance between features within a class is minimised [28]. It is commonly used when there are many data points (features) to process, such as in facial recognition or other image recognition applications that require the extraction of many features. In this study, it was once again implemented using Sci-kit Learn.

#### 3) Logistic Regression (LR)

Logistic regression builds on linear regression analysis, which analyses the relationship between independent predictor variables, and dependent outcome variables, assuming that the relationship between these variables is linear. In the case of logistic regression, the output variables are given to a sigmoid function to convert them to a probability between 0 and 1, thus allowing separation into two classes: those below 0.5 probability and those above [29]. This concept can be extended to multi-class classification, as was the case in the Sci-kit Learn LR algorithm implementation.

#### 4) Artificial Neural Network (ANN)

In contrast to deep neural networks (CNNs) which use 2D layers, ANNs refer to multiple 1D layers of neurons stacked on top of each other for classification of features. They are also commonly known as multi-layer perceptrons, or feed-forward neural networks, and have been successfully used in medical image classification such as classifying CT scans containing lung nodules [30] and skin lesion malignancy [31]. For the current study, the ANN was implemented in Python’s Keras library, using an input layer with the same length as each row of features (560), 5 hidden Dense layers of 550 neurons each, and a Dense-3 layer with softmax activation at the output, as illustrated in Fig. 5. It was trained in a similar fashion to the CNNs that performed the feature extraction, using learning rate reduction with 4 epochs of validation loss patience and early stopping after 8 epochs of no improvement of the validation loss.

**Fig. 5.**
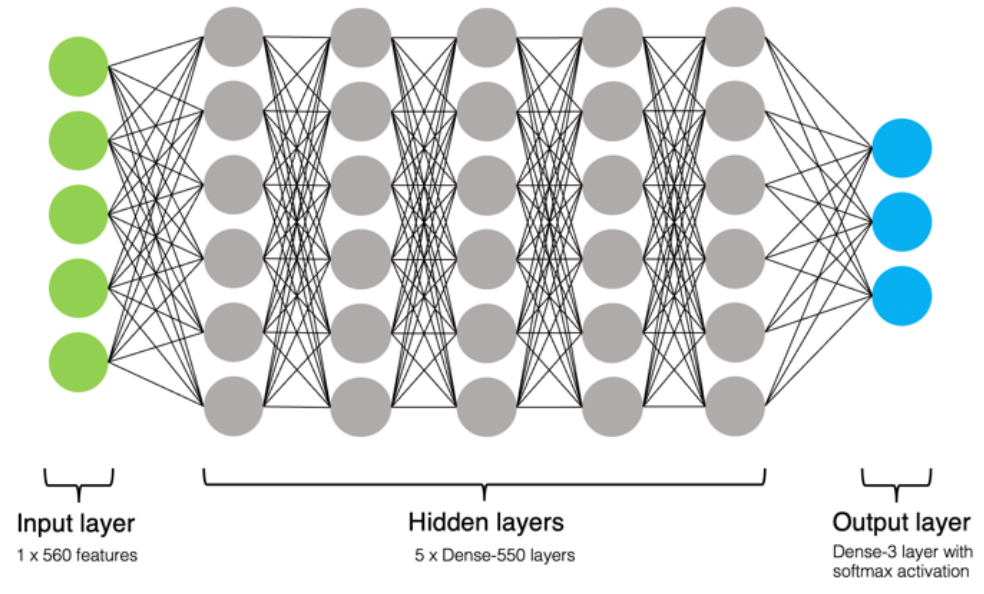
Structure of custom ANN classifier.

Once the above 4 classifiers had been fitted to the features from either the COVID-QU-Ex or the Mostafiz et al. training datasets, the same process was repeated to extract features from their respective test datasets, and then the models were evaluated based on their predictions for each chest x-ray image.

### G. Generalisability of Models

In order to test the generalisability performance of the models when they are trained on different datasets, four variations of dataset training and testing were performed:

1. Training and testing on COVID-QU-Ex dataset.
2. Training and testing on Mostafiz et al. (2022) dataset.
3. Training on COVID-QU-Ex dataset and testing on Mostafiz et al. (2022) dataset.
4. Training on Mostafiz et al. (2022) dataset and testing on COVID-QU-Ex dataset.

The purpose of this experiment is to investigate the influence of dataset size on the degree of overfitting, and the ability of the model to extrapolate to new input images.

### H. Classification Metrics

Several accuracy metrics were computed as outlined in Table 3, where TP/TN, FP/FN are true positive/negative and false positive/negative of class predictions respectively.

**TABLE 3:** Classification Accuracy Metrics

| Metric | Equation |
| --- | --- |
| Accuracy | $\frac{TP + TN}{TP + TN + FP + FN}$ |
| Precision | $\frac{TP}{TP + FP}$ |
| Sensitivity | $\frac{TP}{TP + FN}$ |
| Specificity | $\frac{TN}{TN + FP}$ |
| F1-score | $\frac{2TP}{2TP + FP + FN}$ |

## III. Results

### A. CNN Training Results

The training and validation accuracies during the training of the 3 CNNs are shown in Fig. 6.

**Fig. 6.**
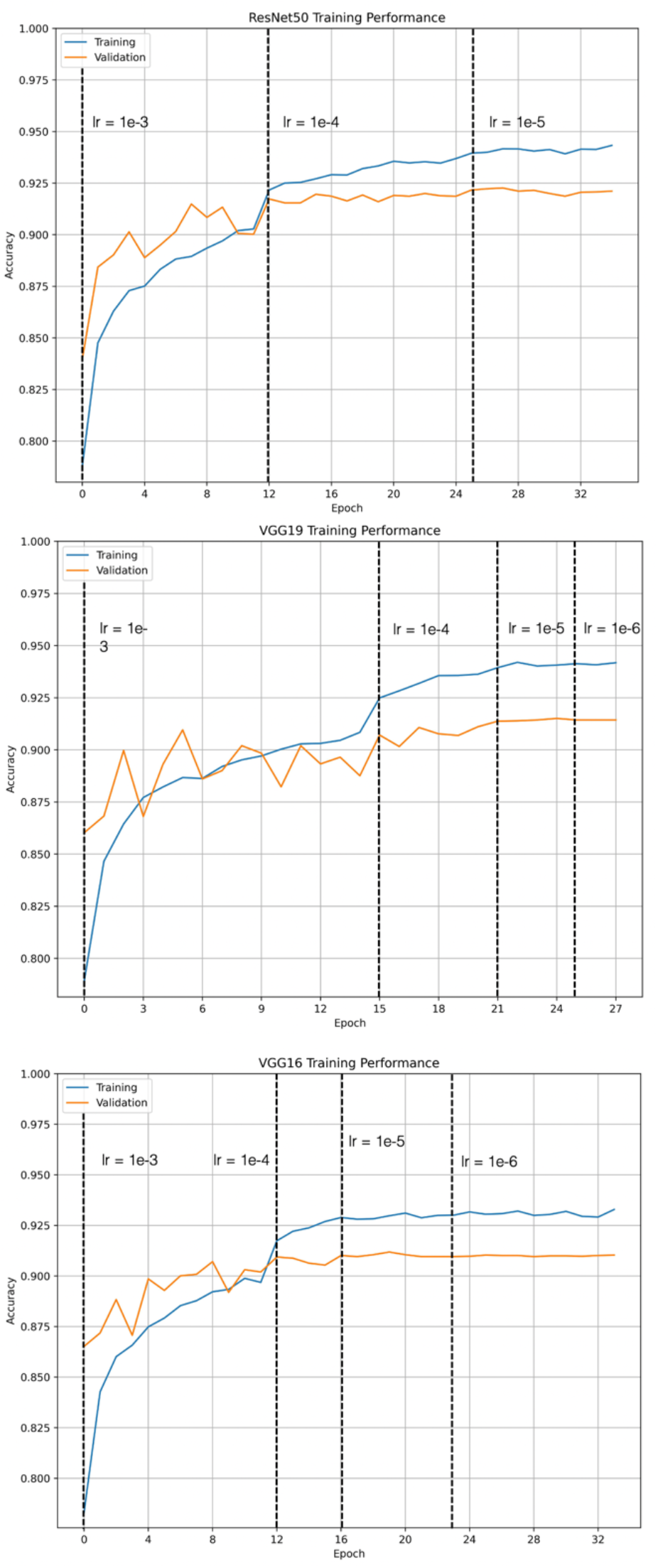
Training curves for ResNet50, VGG16, VGG19 (top to bottom). Vertical dashed lines show when a new learning rate (lr) was applied by the training algorithm.

**Fig. 7.**
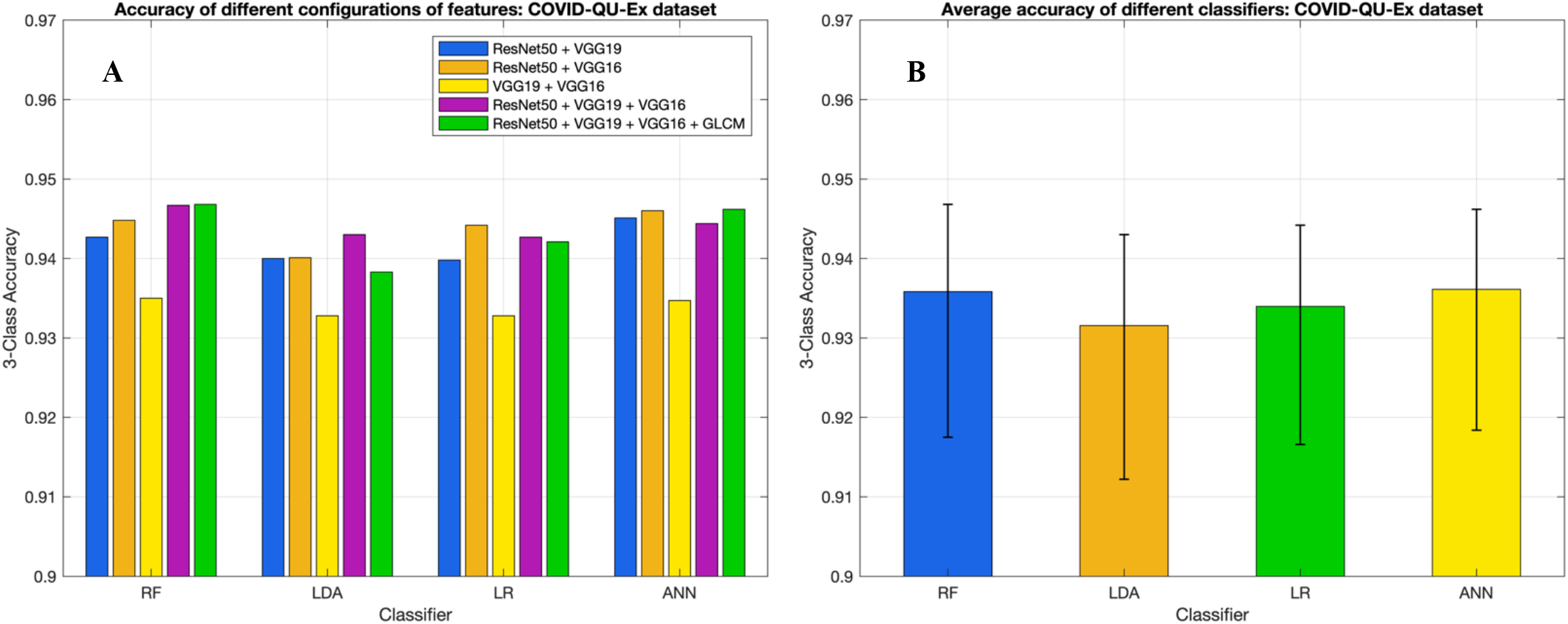
COVID-QU-Ex test dataset classification accuracies for different classifier configurations. **A** - Comparison of accuracies resulting from different feature combinations (features from individual CNNs not shown for clarity). **B** - Average classification accuracy (including individual CNN features) for each type of classifier. Error bars show range of values.

**Fig. 8.**
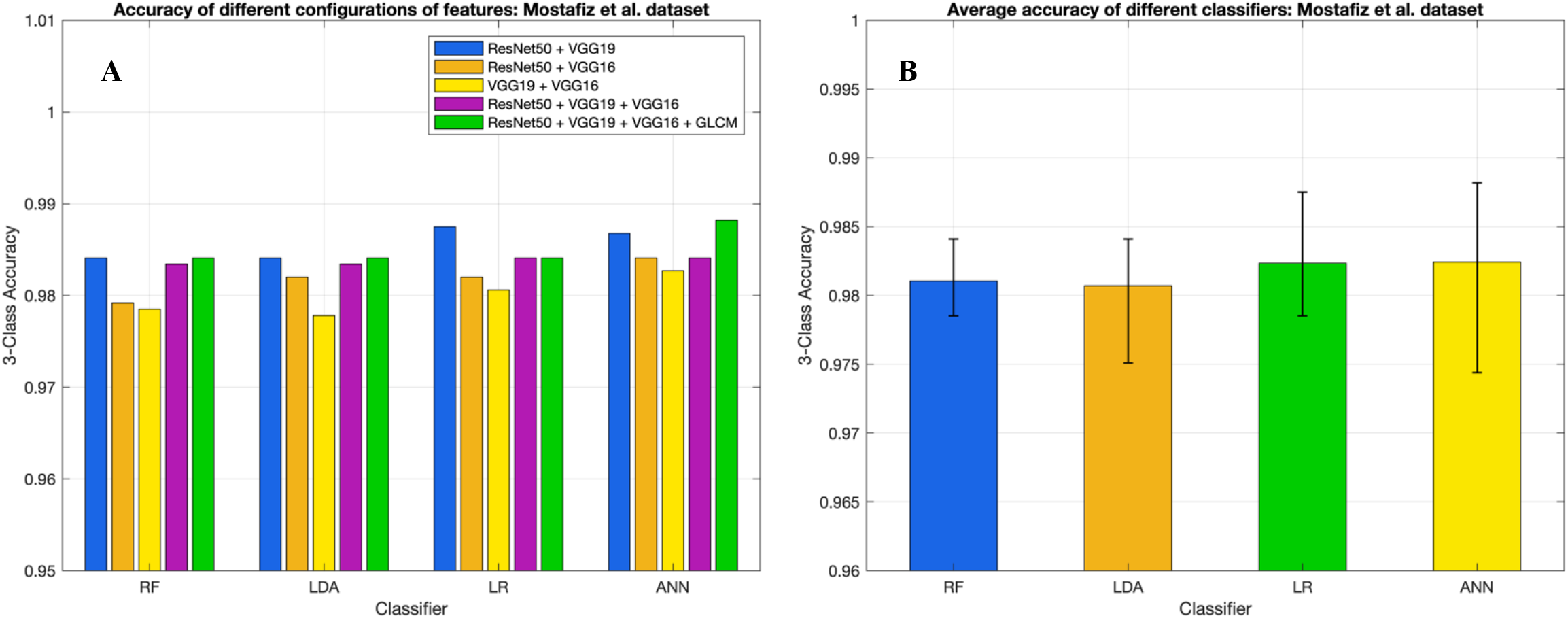
Mostafiz et al. test dataset classification accuracies for different classifier configurations. **A** - Comparison of accuracies resulting from different feature combinations (features from individual CNNs not shown for clarity). **B** - Average classification accuracy (including individual CNN features) for each type of classifier. Error bars show range of values.

The benefits of using LR reduction and early stopping during the network training are clear: Table 4 shows that all 3 CNNs saw an improvement in test dataset accuracy after the modified training regime. In addition, these better test accuracies were achieved in fewer epochs: 35, 28 and 34 for ResNet50, VGG19 and VGG16 respectively, as per Fig. 6.

**TABLE 4:** 3-Class CNN test accuracies with and without LR reduction and early stopping

| CNN | Simple training (40 epochs) | LR reduction + early stopping |
| --- | --- | --- |
| ResNet50 | 0.9184 | 0.9337 |
| VGG19 | 0.9123 | 0.9189 |
| VGG16 | 0.8991 | 0.9231 |

Similar results occurred for the training of the same CNNs on the smaller dataset used by Mostafiz et al. as shown in Table 5. In this case, the accuracies were already higher due to the smaller dataset, therefore decreasing the influence of the LR reduction and early stopping. Nevertheless, an improvement was seen for all CNNs.

**TABLE 5:** 3-Class CNN test accuracies with and without LR reduction and early stopping for Mostafiz et al. dataset

| CNN | Simple training (40 epochs) | LR reduction + early stopping |
| --- | --- | --- |
| ResNet50 | 0.9785 | 0.9806 |
| VGG19 | 0.9674 | 0.9709 |
| VGG16 | 0.9729 | 0.9757 |

### B. Results for Different Classifiers

The test image classification results for each dataset are shown in Table 6 and Table 7. Accuracy refers to the 3-class accuracy for distinguishing between COVID-19, Non-COVID pneumonia and Normal chest x-rays. The other metrics (precision, sensitivity, specificity and F1-score) typically correspond to a single class in a dataset, but in this case they are macro averages of the metrics obtained for each of the 3 classes.

**TABLE 6:** Classification metrics for different combinations of input features: COVID-QU-Ex dataset.

| Classifier and metrics | ResNet50 | VGG19 | VGG16 | ResNet50 + VGG19 | ResNet50 + VGG16 | VGG19 + VGG16 | ResNet50 + VGG19 + VGG16 | ResNet50 + VGG19 + VGG16 + GLCM |
| --- | --- | --- | --- | --- | --- | --- | --- | --- |
| **Random Forest** |  |  |  |  |  |  |  |  |
| Accuracy | **0.9307** | **0.9175** | **0.9225** | **0.9427** | **0.9448** | **0.9350** | **0.9467** | **0.9468** |
| Precision | 0.9304 | 0.9172 | 0.9221 | 0.9425 | 0.9445 | 0.9348 | 0.9463 | 0.9465 |
| Sensitivity | 0.9302 | 0.9170 | 0.9221 | 0.9422 | 0.9445 | 0.9346 | 0.9461 | 0.9463 |
| Specificity | 0.9655 | 0.9589 | 0.9614 | 0.9715 | 0.9726 | 0.9676 | 0.9735 | 0.9735 |
| F1-score | 0.9303 | 0.9171 | 0.9221 | 0.9423 | 0.9445 | 0.9346 | 0.9462 | 0.9464 |
| **LDA** |  |  |  |  |  |  |  |  |
| Accuracy | **0.9268** | **0.9122** | **0.9193** | **0.9400** | **0.9401** | **0.9328** | **0.9430** | **0.9383** |
| Precision | 0.9265 | 0.9119 | 0.9189 | 0.9397 | 0.9398 | 0.9327 | 0.9427 | 0.9386 |
| Sensitivity | 0.9264 | 0.9117 | 0.9189 | 0.9396 | 0.9397 | 0.9325 | 0.9426 | 0.9379 |
| Specificity | 0.9635 | 0.9563 | 0.9598 | 0.9701 | 0.9702 | 0.9666 | 0.9716 | 0.9693 |
| F1-score | 0.9264 | 0.9118 | 0.9189 | 0.9396 | 0.9397 | 0.9325 | 0.9426 | 0.9380 |
| **LR** |  |  |  |  |  |  |  |  |
| Accuracy | **0.9304** | **0.9166** | **0.9231** | **0.9398** | **0.9442** | **0.9328** | **0.9427** | **0.9421** |
| Precision | 0.9301 | 0.9165 | 0.9227 | 0.9395 | 0.9437 | 0.9327 | 0.9424 | 0.9418 |
| Sensitivity | 0.9300 | 0.9163 | 0.9227 | 0.9393 | 0.9437 | 0.9325 | 0.9423 | 0.9416 |
| Specificity | 0.9654 | 0.9585 | 0.9617 | 0.9701 | 0.9722 | 0.9666 | 0.9715 | 0.9712 |
| F1-score | 0.9300 | 0.9163 | 0.9227 | 0.9394 | 0.9437 | 0.9325 | 0.9423 | 0.9417 |
| **ANN** |  |  |  |  |  |  |  |  |
| Accuracy | **0.9299** | **0.9184** | **0.9243** | **0.9451** | **0.9460** | **0.9347** | **0.9444** | **0.9462** |
| Precision | 0.9298 | 0.9183 | 0.9240 | 0.9448 | 0.9471 | 0.9347 | 0.9443 | 0.9460 |
| Sensitivity | 0.9295 | 0.9178 | 0.9241 | 0.9447 | 0.9472 | 0.9343 | 0.9440 | 0.9457 |
| Specificity | 0.9651 | 0.9594 | 0.9623 | 0.9727 | 0.9739 | 0.9675 | 0.9723 | 0.9732 |
| F1-score | 0.9295 | 0.9180 | 0.9239 | 0.9447 | 0.9471 | 0.9344 | 0.9440 | 0.9458 |

**TABLE 7:** Classification metrics for different combinations of input features: Mostafiz et al. DATASET.

| Classifier and metrics | ResNet50 | VGG19 | VGG16 | ResNet50 + VGG19 | ResNet50 + VGG16 | VGG19 + VGG16 | ResNet50 + VGG19 + VGG16 | ResNet50 + VGG19 + VGG16 + GLCM |
| --- | --- | --- | --- | --- | --- | --- | --- | --- |
| **Random Forest** |  |  |  |  |  |  |  |  |
| Accuracy | **0.9813** | **0.9785** | **0.9792** | **0.9841** | **0.9792** | **0.9785** | **0.9834** | **0.9841** |
| Precision | 0.9798 | 0.9794 | 0.9801 | 0.9827 | 0.9770 | 0.9792 | 0.9820 | 0.9827 |
| Sensitivity | 0.9844 | 0.9814 | 0.9786 | 0.9862 | 0.9831 | 0.9817 | 0.9858 | 0.9862 |
| Specificity | 0.9897 | 0.9879 | 0.9883 | 0.9911 | 0.9888 | 0.9881 | 0.9907 | 0.9911 |
| F1-score | 0.9821 | 0.9804 | 0.9794 | 0.9844 | 0.9800 | 0.9804 | 0.9838 | 0.9844 |
| **LDA** |  |  |  |  |  |  |  |  |
| Accuracy | **0.9827** | **0.9764** | **0.9751** | **0.9841** | **0.9820** | **0.9778** | **0.9834** | **0.9841** |
| Precision | 0.9813 | 0.9741 | 0.9779 | 0.9825 | 0.9805 | 0.9760 | 0.9820 | 0.9831 |
| Sensitivity | 0.9860 | 0.9804 | 0.9762 | 0.9872 | 0.9855 | 0.9816 | 0.9864 | 0.9872 |
| Specificity | 0.9904 | 0.9874 | 0.9848 | 0.9912 | 0.9901 | 0.9882 | 0.9907 | 0.9912 |
| F1-score | 0.9836 | 0.9772 | 0.9770 | 0.9848 | 0.9830 | 0.9787 | 0.9842 | 0.9851 |
| **LR** |  |  |  |  |  |  |  |  |
| Accuracy | **0.9820** | **0.9785** | **0.9799** | **0.9875** | **0.9820** | **0.9806** | **0.9841** | **0.9841** |
| Precision | 0.9805 | 0.9792 | 0.9829 | 0.9877 | 0.9799 | 0.9810 | 0.9836 | 0.9836 |
| Sensitivity | 0.9855 | 0.9821 | 0.9802 | 0.9890 | 0.9870 | 0.9827 | 0.9881 | 0.9881 |
| Specificity | 0.9901 | 0.9881 | 0.9875 | 0.9929 | 0.9908 | 0.9892 | 0.9915 | 0.9915 |
| F1-score | 0.9830 | 0.9806 | 0.9815 | 0.9884 | 0.9834 | 0.9819 | 0.9858 | 0.9858 |
| **ANN** |  |  |  |  |  |  |  |  |
| Accuracy | **0.9806** | **0.9785** | **0.9744** | **0.9868** | **0.9841** | **0.9827** | **0.9841** | **0.9882** |
| Precision | 0.9787 | 0.9777 | 0.9719 | 0.9854 | 0.9839 | 0.9834 | 0.9839 | 0.9872 |
| Sensitivity | 0.9837 | 0.9832 | 0.9793 | 0.9895 | 0.9856 | 0.9849 | 0.9861 | 0.9916 |
| Specificity | 0.9893 | 0.9891 | 0.9870 | 0.9928 | 0.9907 | 0.9905 | 0.9914 | 0.9940 |
| F1-score | 0.9812 | 0.9802 | 0.9753 | 0.9875 | 0.9847 | 0.9841 | 0.9850 | 0.9893 |

For the COVID-QU-Ex dataset, the Random Forest classifier had the best maximum performance at 94.68% accuracy on a test dataset of 6588 images. However, across all of the feature combinations, the ANN classifier had a slightly better performance on average, at 93.61% compared to 93.58% for the RF classifier. In terms of feature combinations, the feature combination of VGG19 and VGG16 features gave consistently the worst performance, whilst the highest recorded accuracy came from the combination of all features and passed to the RF classifier.

For the Mostafiz et. al dataset, RF, LDA and LR all obtained the same accuracies on the combination of all features, while the ANN outperformed them, achieving a maximum accuracy of 98.82% on the test dataset of 1443 images. On average, the best feature combination was ResNet50 with VGG19 features at 98.56% average accuracy across the different classifiers, however combining all feature types was only a slightly smaller average performance at 98.51%.

### A. COVID-19 Detection Performance

One of the primary aims of this model is to improve on the performance and sensitivity of the PCR test and the general triaging process for patients with COVID-19 pneumonia. This therefore warrants an examination of the binary classification accuracy and the specific sensitivity to COVID-19. Fig. 9 and Fig. 10 show the confusion matrices for a 3-class classification as well as for a binary classification that only considers whether COVID-19 was detected or not. They correspond to the models that performed the best for each dataset, being the RF classifier with all features for the larger COVID-QU-Ex dataset, and the ANN classifier with all features for the smaller dataset from Mostafiz et al. Classification metrics for each are shown in Table 8 and Table 9.

**TABLE 8:** Metrics for RF classifier with COVID-QU-Ex Dataset

| Metric | Performance |
| --- | --- |
| 3-Class Accuracy | 0.9468 |
| Binary Accuracy | 0.9843 |
| Sensitivity to COVID-19 | 0.9713 |

**TABLE 9:** Metrics for ANN classifier with Mostfiz et al. Dataset

| Metric | Performance |
| --- | --- |
| 3-Class Accuracy | 0.9882 |
| Binary Accuracy | 0.9986 |
| Sensitivity to COVID-19 | 1.0 |

**Fig. 9.**
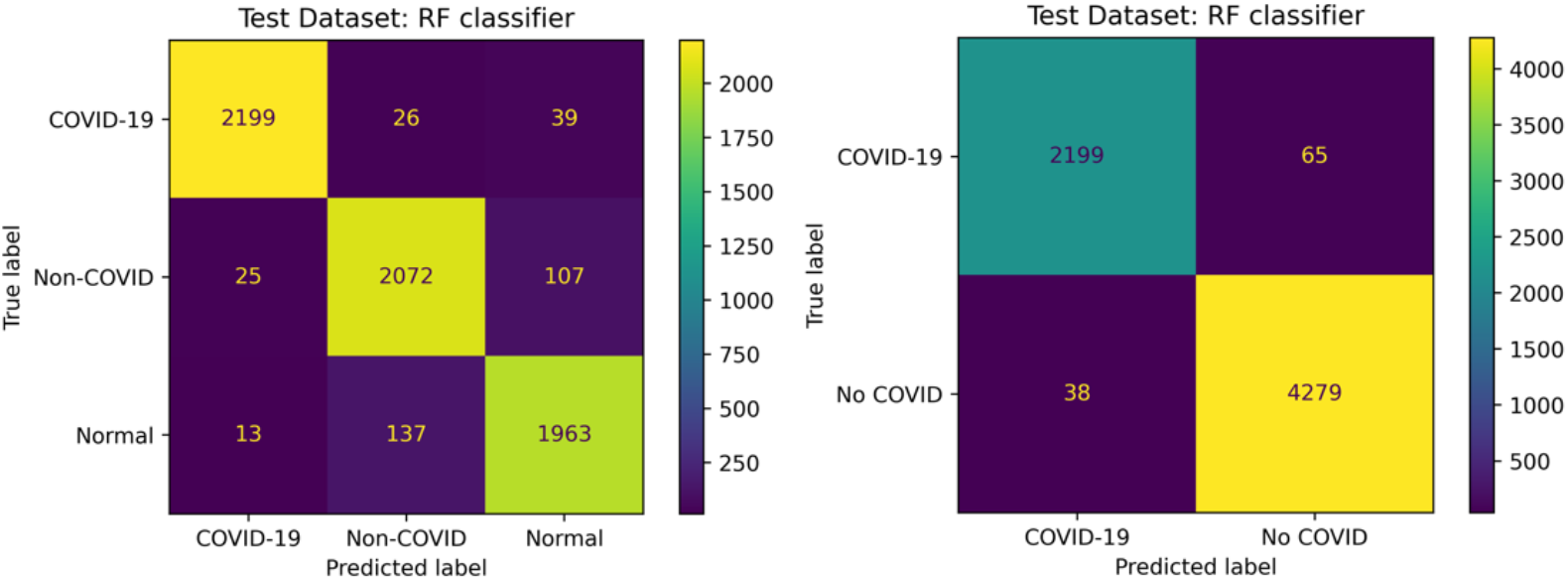
3-Class (left) and binary (right) classification confusion matrices for highest performing model in COVID-QU-Ex dataset testing: RF classifier with all features.

**Fig. 10.**
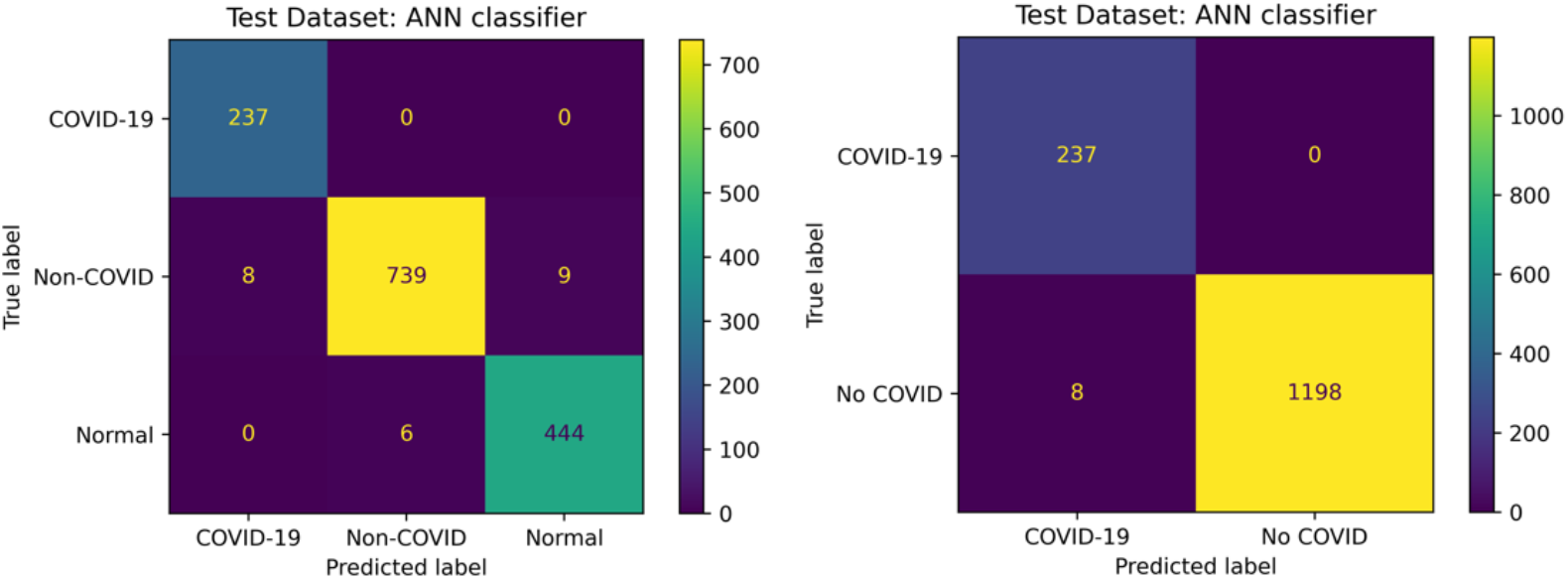
3-Class (left) and binary (right) classification confusion matrices for highest performing model in Mostafiz et al. dataset testing-ANN classifier with all features.

### D. Generalisability of Models to Foreign Image Data

The models clearly perform well on their own respective test datasets. In reality, however, a robust model used clinically would require that it generalise well to any input chest x-ray image, not just to the test partition of the dataset that it was trained on. To examine the performance on images external to the training dataset of the model (foreign data), a cross-dataset testing procedure was performed. This involved testing the model that was trained on the COVID-QU-Ex dataset using the dataset from Mostafiz et al., and vice versa. To ensure fairness, the COVID-QU-Ex test dataset was modified to match the numbers of chest x-rays in each class of the Mostafiz et al. test dataset. This was done by randomly selecting 237 COVID-19, 756 Non-COVID, and 450 normal chest x-rays from the COVID-QU-Ex test dataset. All 4 types of end classifier were examined, with the results shown in Fig. 11 and Fig. 12.

**Fig. 11.**
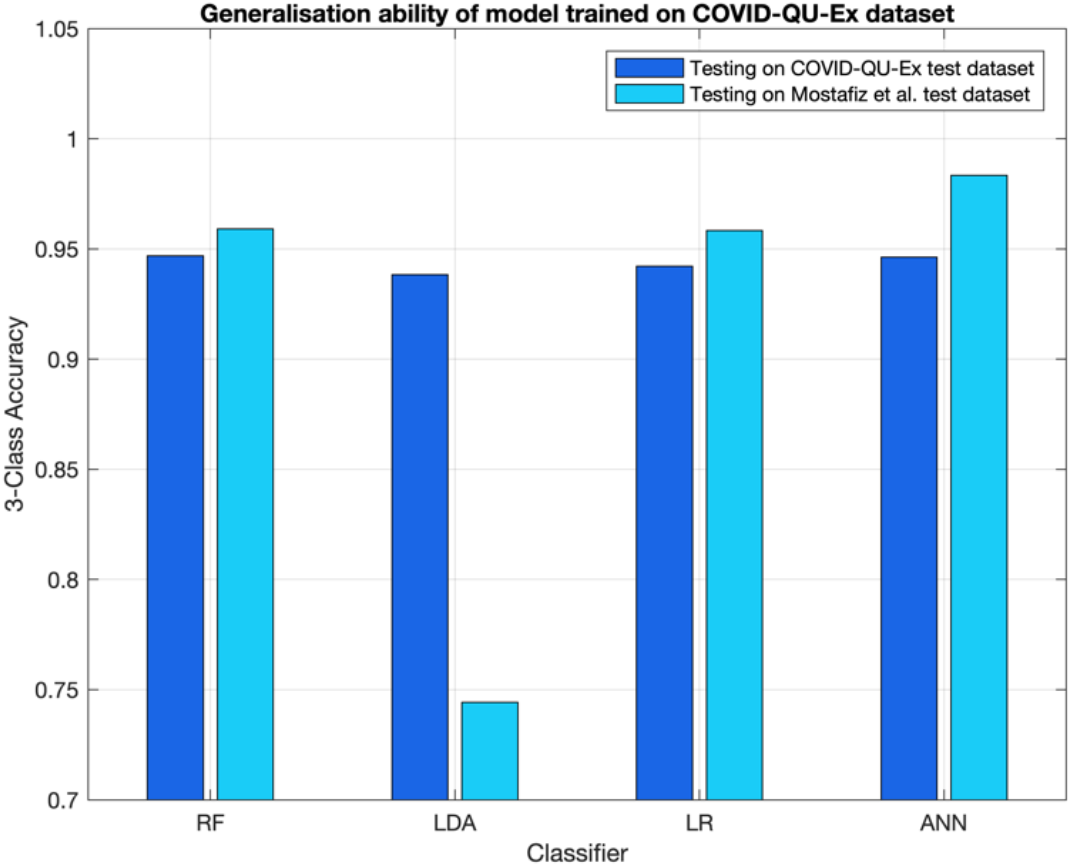
Results of training model on large dataset and testing on foreign dataset.

**Fig. 12.**
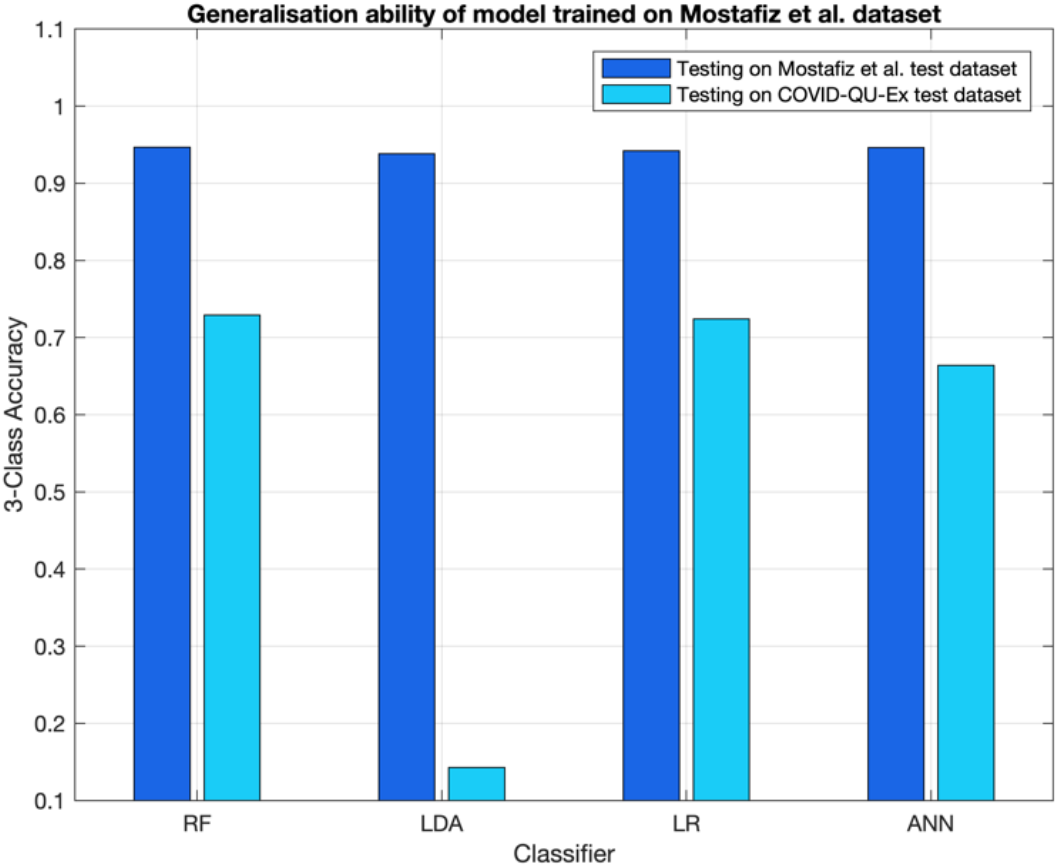
Results of training model on small dataset and testing on foreign dataset.

It is apparent in Fig. 11 that, other than for the LDA classifier, the generalisation of the model trained on the larger dataset is excellent, achieving accuracies even higher than for its own test dataset. On the other hand, evidently the LDA classifier is severely prone to overfitting, not generalising well to new data. This was also clear in Fig. 12 for the model trained on the smaller Mostafiz et al. dataset and tested on data from COVID-QU-Ex. In contrast to Fig. 11, however, Fig. 12 shows that the other classifiers also did not generalise well for this dataset, achieving accuracies of only around 70% for RF, LR and ANN. This points to an issue with the feature extraction CNNs poorly extracting features from the foreign images. To summarise, the results reaffirm that training on a larger (>20,000 image) dataset allows the end model to better generalise to new data than training on a smaller (∼4000 image) dataset.

## IV. Discussion

The results of training the CNNs show that it is possible to improve their test dataset accuracy by strategically lowering the learning rate and by using early stopping. Learning rate reduction is also known as learning rate scheduling, and has a significant influence on gradient descent in the training process. Having a relatively large learning rate when training begins allows a rough and rapid estimation of the model minimum loss, with further decreases in learning rate tuning the model weights with finer and finer steps until the loss converges to a global minimum [32]. This is analogous to first using a coarse focus followed by a fine focus to visualise an object under a microscope with a high magnification. Continuously using the same high learning rate throughout the training process makes it far more difficult for the weights to converge to their ideal values since their values are shifted by far greater amounts, just like only using only using coarse focus on a microscope. This can cause the model to converge at local minima instead [33]. Conversely, only using a low learning rate will substantially increase the computation time, and likewise may get stuck at local minima. Using strategic learning rate reduction, test accuracies were improved by 1.53% on average, and required less training epochs, reducing the computational load.

The results of the classification exemplify the benefits of ensemble techniques in medical image classification for improving the accuracies obtained via CNN classification. For the COVID-QU-Ex dataset, the mean accuracy for classification of features from individual CNNs was 0.9236 for the RF classifier. However, the combination of features from each CNN, GLCM features and classification using traditional machine learning classifiers yielded a maximum 3-class accuracy of 0.9468 with the RF classifier, a substantial improvement. The benefits resulting from such ensemble CNN approaches have been documented in other studies. Togacar et al. (2020) used a similar CNN feature concatenation approach for pneumonia detection in chest x-rays, and attained a binary accuracy about 2.7% higher than for their individual CNNs [10]. The approach appears to be able to be extended to other specific diseases such as tuberculosis detection, as demonstrated by Hooda et al. (2019), who saw a 5.5% increase in TB detection accuracy when combining the features extracted by AlexNet, GoogleNet and ResNet34 [34].

The results in Fig. 9 and Fig. 10 show that, for both datasets, any confusion mostly resided in distinguishing between Non-COVID pneumonia and Normal chest x-rays and not significantly between either of these and the COVID-19 chest x-rays. This means that the binary classification for COVID-19 or not COVID-19 was in both cases very good, at 98.43% accuracy for the large dataset and 99.86% for the smaller dataset. The binary COVID-19 detection accuracy was, in fact, slightly improved over that obtained by Mostafiz et al. who achieved 99.45% [6]. The sensitivity to COVID-19 was similarly high, at 97.13% for the large dataset and 100% in the case of the smaller dataset. Both instances perform better than a PCR, which is on average about 90.7% sensitive to COVID-19 [1]. Fig. 11 shows that as long as the model is trained on sufficient data, these accurate metrics can be maintained when applying the model to new data, allowing it to adequately be used as a clinical diagnostic tool.

There have been numerous research articles presenting COVID-19 chest x-ray classification models trained on small datasets of only a few hundred to a couple thousand images, some of which document very high (>98) accuracies [3]-[9]. The significance of the present study is that it shows that high accuracy on small datasets does not mean that the model is generalisable and robust to other datasets, which is the overall aim of developing such models in the first place. Generalisability of machine learning models is especially critical in a clinical environment, where, between hospitals, there may be differences in medical image acquisition systems, patient demographics, and professional training [35]. It is well understood that increasing dataset size improves the ability of CNNs and other machine learning classifiers to fit input data and reduce dataset overfitting [36][37]. The cleaned COVID-QU-Ex dataset used for training contained 11380 chest x-rays with COVID-19, 11048 with Non-COVID pneumonia and 10529 with no condition. Due to the large number and, therefore, variety of images, the CNNs learned more general features during training, and were therefore able to generalise very well when exposed to foreign chest x-ray images from Mostafiz et al, obtaining an average 96.7% accuracy across different classifiers (excluding LDA classifier outlier). On the other hand, the Mostafiz et al. dataset contained only 790 cases of COVID-19, 2519 Non-COVID pneumonia, and 1500 with no condition. Consequently, the CNNs learned to extract features that were specific to this dataset very accurately, but generalised poorly when given the COVID-QU-Ex images. The models obtained an average accuracy of 70.57% across the different classifiers (excluding LDA classifier outlier). The consequence of this result is that training dataset size has a direct impact on the accuracy of predictions and must be considered when attempting to develop clinically relevant and robust automatic classification models.

For both cross-dataset tests, the LDA classifier performed poorly, clearly a sign of overfitting the image features from dataset on which it was trained. Unlike for the similar Principle Component Analysis (PCA) where insignificant dimensions are ignored, they are included in the LDA process, causing the model to fit specific features rather than general ones [28][38]. This may make this particular classifier unsuitable in scenarios such as medical image classification where there are typically a high number of input features, and there may be differences in x-ray acquisition systems that can introduce variability in the images. On the other hand, RF, LR and ANN classifiers appear to generalise well to new features. In particular, the COVID-QU-Ex-trained ANN classifier achieved an outlying 98.34% accuracy on the unseen Mostafiz et al. dataset, suggesting its superior usage when attempting to classify new chest x-ray images.

## V. Conclusion

This study examined several techniques in medical image deep learning, and elucidated the benefits of combining CNNs for improved classification performance. It first was found that using learning rate reduction/scheduling can reduce CNN training time, yet substantially improve their test dataset classification performance. In a similar fashion, mRMR feature selection reduces the computation time for fitting features to other classifiers whilst preserving relevant image information. The maximum classification accuracy for the COVID-QU-Ex dataset was achieved when extracted features of all of ResNet50, VGG19, VGG16 and GLCM features were combined, at 94.68% with the Random Forest classifier. Detection accuracy and sensitivity to COVID-19 were very high, at 98.43% and 97.13% respectively. These were even higher for the Mostafiz et al. dataset, 99.86% binary accuracy and 100% sensitivity, however it was found that the small number of images caused the model to overfit the data, leading to poor generalisation for all classifier types. It is recommended, therefore, that the creation of new or improved COVID-19 or pneumonia classification models prioritise training with large datasets. It is also recommended to avoid the use of LDA classifiers when giving large numbers of input features, due to its poor generalisation ability in medical image classification.

## Data Availability

All data produced in the present work are contained in the manuscript.

